# Efficacy of face masks, neck gaiters and face shields for reducing the expulsion of simulated cough-generated aerosols

**DOI:** 10.1101/2020.10.05.20207241

**Authors:** William G. Lindsley, Francoise M. Blachere, Brandon F. Law, Donald H. Beezhold, John D. Noti

## Abstract

Face masks are recommended to reduce community transmission of SARS-CoV-2. One of the primary benefits of face masks and other coverings is as source control devices to reduce the expulsion of respiratory aerosols during coughing, breathing, and speaking. Face shields and neck gaiters have been proposed as an alternative to face masks, but information about face shields and neck gaiters as source control devices is limited. We used a cough aerosol simulator with a pliable skin headform to propel small aerosol particles (0 to 7 µm) into different face coverings. An N95 respirator blocked 99% of the cough aerosol, a medical grade procedure mask blocked 59%, a 3-ply cotton cloth face mask blocked 51%, and a polyester neck gaiter blocked 47% as a single layer and 60% when folded into a double layer. In contrast, the face shield blocked 2% of the cough aerosol. Our results suggest that face masks and neck gaiters are preferable to face shields as source control devices for cough aerosols.

## Introduction

SARS-CoV-2, the virus that causes coronavirus disease 2019 (COVID-19), can be transmitted from person-to-person by large respiratory aerosols (airborne liquid droplets and dried particles greater than about 10 µm in diameter) produced by people who are infectious while they are talking, singing, coughing, breathing or sneezing (CDC 2020a; Hamner et al. 2020). Smaller aerosols also are emitted by people during these activities, suggesting that short-range airborne transmission of SARS-CoV-2 might be possible under some circumstances (Anderson et al. 2020; CDC 2020a; Ma et al. 2020; Morawska and Milton 2020). To interrupt this potential transmission route, the Centers for Disease Control and Prevention (CDC), the World Health Organization (WHO), and other public health organizations recommend the wearing of face masks or other face coverings by the general public during the ongoing COVID-19 pandemic (CDC 2020b; c; Edelstein and Ramakrishnan 2020; WHO 2020). One of the primary benefits of face coverings is to act as source control devices to reduce the expulsion of aerosols containing the virus from people who are infectious during coughing, breathing, and speaking. Source control devices are intended to protect other people from infectious aerosols emitted by the wearer, as compared with personal protective equipment such as N95 respirators which are primarily intended to protect the wearer. Studies using manikins (Lai et al. 2012; Patel et al. 2016) and patients with respiratory infections (Leung et al. 2020; Milton et al. 2013) have shown that wearing medical face masks can reduce the dispersion of potentially infectious aerosols from patients. Two studies in which face masks were required for visitors and healthcare workers interacting with patients in bone marrow transplant centers found a reduction in respiratory viral infections among patients (Sokol et al. 2016; Sung et al. 2016). Studies of cloth face masks have suggested that they also can be effective at reducing the release of respiratory aerosols into the environment (Asadi et al. 2020; Davies et al. 2013; Konda et al. 2020).

Unfortunately, the use of face masks and other face coverings by the general public can present challenges. People often dislike wearing masks, and compliance can be low and inconsistent (Longtin et al. 2009). Mask wearers may repeatedly don, doff and adjust face masks, which can contaminate the hands and potentially lead to disease transmission, especially when the masks are reused (Brady et al. 2017; Casanova et al. 2008). For cloth masks, the filtration efficiency and air flow resistance of different textiles varies widely (Konda et al. 2020; Teesing et al. 2020; Wilson et al. 2020). Alternative face coverings such as neck gaiters (an elastic fabric tube that fits snugly around the head and neck) are commonly used, but information about their performance as source control devices is limited. Factors such as how well the mask fits the face and the coverage provided by a mask can have a substantial impact on the effectiveness of face masks (Davies et al. 2013; Lawrence et al. 2006). Comparisons of face coverings have found substantial differences in the ability of different types of these devices to reduce the release of respiratory aerosols (Asadi et al. 2020; Davies et al. 2013).

An opinion article in JAMA proposed that face shields would be more effective than face masks at reducing community disease transmission, in large part because the authors felt that face shields were more comfortable and thus that they were more likely to be widely adopted by the public (Perencevich et al. 2020). A previous study by our group of face shields used as personal protective devices showed that face shields protect the wearer from large cough aerosols directed at the face but are much less effective against smaller aerosols which were able to flow around the edges of the shield and be inhaled (Lindsley et al. 2014). However, very little work has been done examining face shields as source control devices. Two qualitative flow visualization studies of face shields and masks found that, although face shields deflected the air flow from the mouth, they did not stop aerosol particles from traveling around the face shield and entering the environment (Verma et al. 2020; Viola et al. 2020). Beyond these studies, quantitative data on the efficacy of face shields for source control are lacking.

The objective of our study was to conduct a quantitative comparison of the efficacy of an N95 respirator, a medical procedure mask, a commercial 3-ply cloth face mask, a single and double layer fabric neck gaiter, and a commercial disposable face shield as source control devices to reduce the expulsion of small cough-generated aerosol particles into the environment. Our results provide more information about the effectiveness of different types of source control devices and will help the public health community make recommendations about the best ways to use these devices to help reduce the spread of COVID-19.

## Materials and Methods

### Experimental Design

In our experiments, a cough aerosol simulator propelled a test aerosol through a headform into a collection chamber (Figure 1), and the amount of aerosol in the collection chamber was measured in each of six size fractions. The collection efficiency of each face mask, neck gaiter, or face shield was determined by comparing the amount of aerosol that was collected from the chamber with and without the device. Our test method was similar to the modified Greene and Vesley method used to test medical masks (Quesnel 1975), with the human test subject replaced by the cough aerosol simulator.

**Figure 1:**
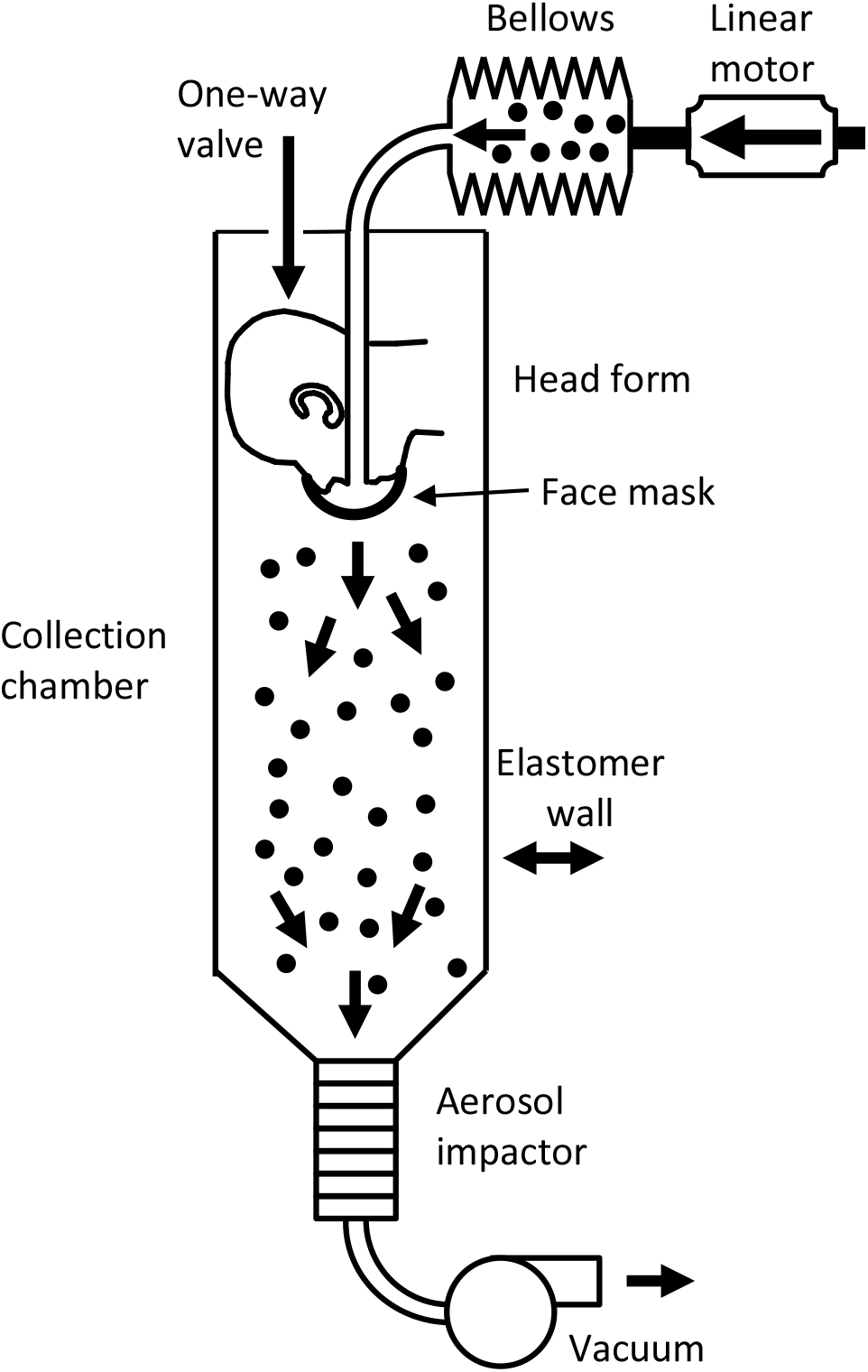
Cough aerosol simulator system for source control measurements. The system consists of an aerosol generation system, a bellows and linear motor to produce the simulated cough, a pliable skin head form on which the face mask, neck gaiter or face shield is placed, a 105 liter collection chamber into which the aerosol is coughed, and an Andersen impactor to separate the aerosol particles by size and collect them. More information about the cough aerosol simulator is provided in the supplemental online materials.

### Cough aerosol simulator

The cough aerosol simulator is a modified version of the NIOSH cough aerosol simulator described previously (Lindsley et al. 2019; Lindsley et al. 2014; Lindsley et al. 2013). The experimental cough aerosol was generated by nebulizing a solution of 14% KCl and 0.4% sodium fluorescein using a single-jet Collison nebulizer (BGI, Butler, NJ) at 103 kPa (15 lbs./in^2^), passing the aerosol through a diffusion drier (Model 3062, TSI, Shoreview, MN), and mixing it with 10 L/min of dry filtered air. The test aerosol was loaded into an elastomeric bellows, and the cough airflow was produced by a computer-controlled linear motor that compresses the bellows. The cough aerosol was expelled through the mouth of a headform into a collection chamber. The headform used in the study has pliable skin that mimics the elastic properties of human skin in order to create a realistic simulation of how each face covering or shield would fit a human face (Bergman et al. 2014).

### Source control devices

The source control devices tested were an N95 medical respirator (3M model 1860), a medical grade (ASTM Level 3) procedure mask with ear loops (Kimberly-Clark model 47107), a cloth face mask with 3 layers of cotton fabric and ear loops (Hanes Defender), a fabric neck gaiter (FKGIONG Sun UV Protection Neck Gaiter, 95% polyester, 5% Spandex) and a disposable face shield (Fisher Scientific # 19-181-600A). The neck gaiter was tested both as a single layer of fabric and doubled over to provide two layers of fabric. The masks and respirator were not equipped with exhalation valves. The face shield was 25 cm tall and extended from the forehead of the headform to 3 cm below the chin and around the side to 3 cm before the front of the ear. Photographs of the source control devices on the headform are shown in the supplemental online materials.

### Mask fit test

For the experiments, either no device, a face mask, a neck gaiter, or a face shield were placed on the head form. Each device was used for two consecutive tests. For face masks and gaiters, a respirator fit test was performed using a PortaCount (TSI). The fit factor is a measure of the protection against airborne particles that is provided by a respiratory protective device. It is defined as the ratio of the aerosol concentration outside the respiratory protective device to the aerosol concentration inside the device (i.e., the aerosol concentration that is inhaled by the wearer). For example, a fit factor of 10 means that the ambient aerosol concentration is 10 times higher than the concentration inside the mask, and that the mask is therefore filtering out 90% of the ambient aerosol.

### Aerosol collection and analysis

After placing the device on the headform and performing the fit test, the system was sealed. The test aerosol was then generated and propelled with a simulated cough through the headform and into the collection chamber. The Andersen impactor at the bottom of the collection chamber collected the aerosol particles that traveled through or around the device for 20 minutes after each cough. The Andersen impactor operates at a flow rate of 28.3 liters/minute and has six collection stages and a filter that separate the aerosol particles into seven size fractions based on the aerodynamic diameter of the particles: <0.6 µm; 0.6-1.1 µm; 1.1-2.1 µm; 2.1-3.3 µm; 3.3-4.7 µm; 4.7-7.0 µm; and >7 µm. Because the amount of aerosol in the largest size fraction was small and because of possible losses due to settling of the large aerosol particles, data for the largest size fraction was not included in the analysis. The impactor collection plates were coated with a solution of glycerol and Brij 35 to prevent particles from bouncing off the plates during collection (Mitchell 2003). After aerosol collection was completed, the impactor plates were rinsed with 0.1 M Tris solution and the fluorescence of the solution was measured using a fluorometer (SpectraMax M4, Molecular Devices). The complete experimental protocol is given in the supplemental online materials.

### Statistical Analysis

The performance of each device was evaluated by comparing the total mass of the aerosol particles from a single cough that passed through or around the device and was collected by the Andersen impactor. The results were evaluated using a one-way ANOVA and multiple comparisons among the different devices and the control experiments without a device were conducted using a Tukey-Kramer test. To control for variations in the amount of aerosol in each cough, a sample of each cough aerosol was collected from the bellows prior to coughing and used to normalize the aerosol mass collection results for each experiment.

## Results

The cough aerosol simulator provides a cough with a controlled cough airflow rate containing a test aerosol with a consistent aerosol size distribution. The simulator allows for a direct quantitative comparison of the ability of different types of source control devices to block the expulsion of simulated cough aerosol particles of different sizes into the environment. The flow rate of the simulated cough used in our experiments was based on cough flow profiles recorded from influenza patients and had a volume of 4.2 L with a peak flow rate of 11 L/s (Lindsley et al. 2013). The cough aerosol collected from the control experiments without a face covering had a mass median aerodynamic diameter of 1.3 µm, a geometric standard deviation of 2.3 and a total aerosol mass of 505 µg (standard deviation 69).

For our study, we tested the collection efficiencies (that is, the fraction of the cough aerosol that was blocked) of a medical grade procedure mask, a cotton cloth face mask, a polyester neck gaiter, an N95 medical respirator and a disposable face shield. These source control devices were chosen to provide representative samples of the different types of face coverings and face shields that are in common use during the pandemic. Neck gaiters are typically worn either as a single layer of fabric over the mouth and nose or doubled over to provide two layers of fabric; for our experiments, we tested both configurations. The quantity of aerosol particles in six size fractions that were able to travel through or around each source control device are shown in Figure 2. The collection efficiencies of the devices are shown as a function of aerosol size in Figure 3. All the devices showed increased collection efficiencies as the aerosol size increased.

**Figure 2:**
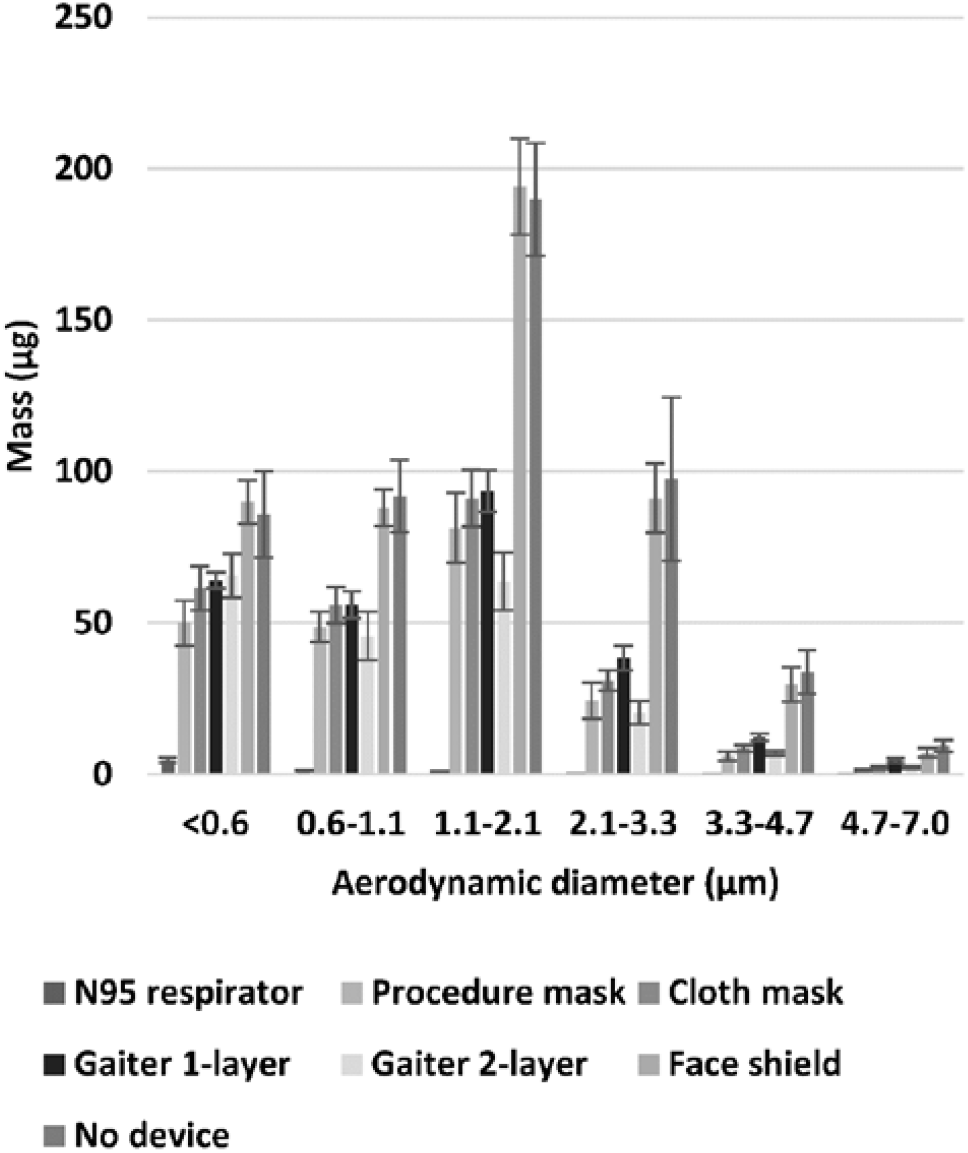
Mass of aerosol collected in each size fraction. The graph shows the amount of simulated respiratory aerosol that was collected from the collection chamber in each aerosol particle size fraction after a single simulated cough. The bars show the mean and standard deviation. A larger color version of this figure is shown in the supplemental online materials.

**Figure 3:**
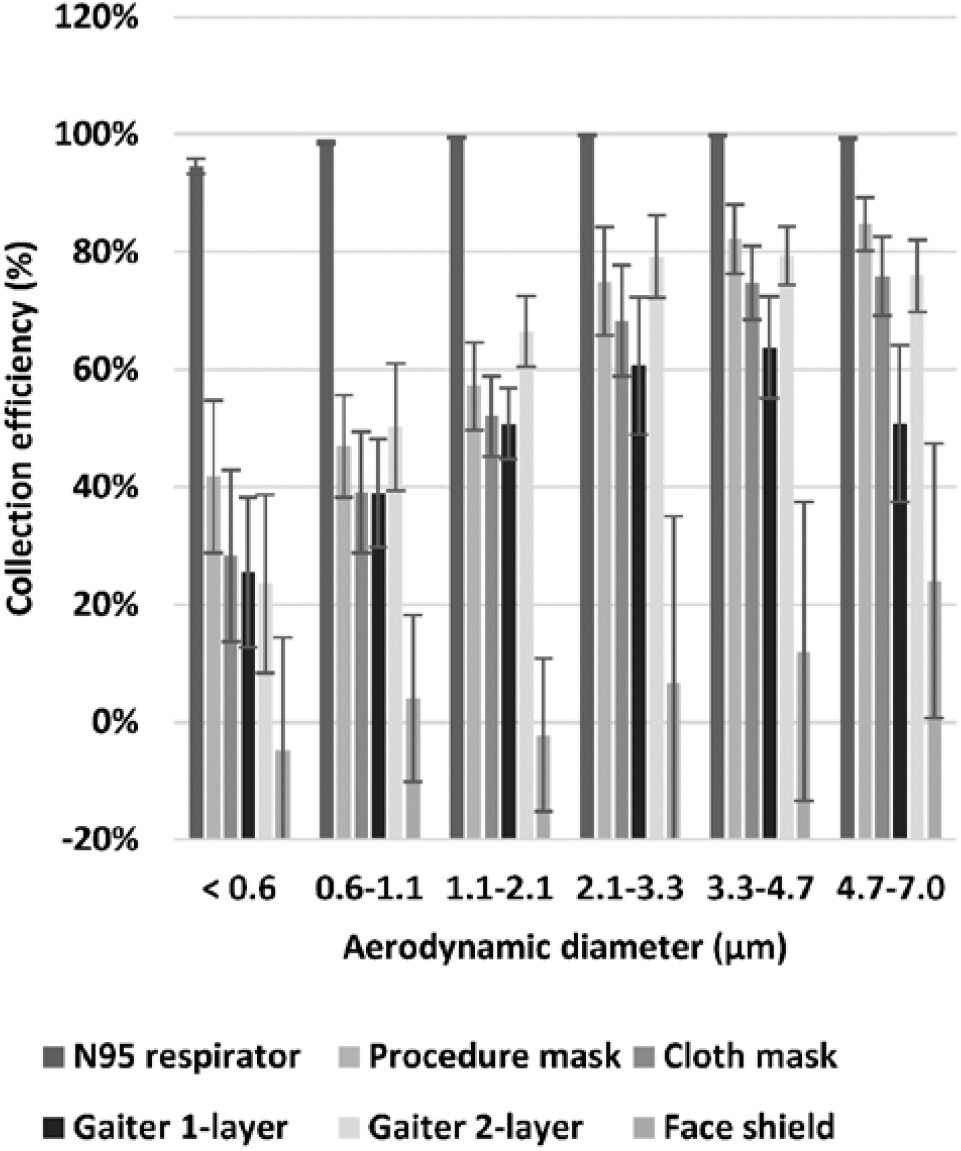
Collection efficiency of face masks, neck gaiter and face shield. The collection efficiency is the percentage of aerosol particles that were blocked by the face mask, neck gaiter or face shield compared with experiments without a device. The plot shows the means and standard deviations of the collection efficiency in each size fraction. A larger version of this figure is shown in color in the supplemental online materials.

On average, the N95 respirator blocked 99% of the total mass of test aerosol from being released into the environment, while the medical procedure mask blocked 59%, the cloth face mask blocked 51%, the single-layer gaiter blocked 47%, the double-layer gaiter blocked 60%, and the face shield blocked 2% of the total aerosol (Table 1). The N95 respirator, procedure mask, cloth mask, and the single-layer and double-layer gaiters all significantly reduced the aerosol emitted into the environment compared with no device (P < 0.0001 for each), but the face shield did not (P = 0.9993). The collection efficiencies of the procedure mask, cloth mask, and the single and double-layer gaiters did not differ significantly from each other, but all blocked cough aerosols significantly better than did the face shield (P <0.0001). The N95 respirator outperformed all the other devices (P < 0.0001) (Table 2).

**Table 1:** Total mass of aerosol expelled into collection chamber and device collection efficiencies. The fit factor, aerosol mass, and collection efficiency are given as mean (standard deviation).

| *Device tested* | *Number of experiments* | *Fit factor* | *Aerosol mass ($\mu\text{g}$)* | *Collection efficiency* |
| --- | --- | --- | --- | --- |
| No device | 12 | n/a | 512 (64) | n/a |
| Procedure mask | 6 | 2.9 (0.5) | 212 (23) | 58.5% (6.9%) |
| Cloth mask | 6 | 1.3 (0.1) | 251 (23) | 50.9% (7.7%) |
| Neck gaiter (single layer) | 6 | 1.7 (0.5) | 270 (18) | 47.2% (7.5%) |
| Neck gaiter (double layer) | 6 | 1.9 (0.4) | 206 (26) | 59.8% (7.2%) |
| Face shield | 6 | n/a | 502 (46) | 1.8% (15.3%) |
| N95 respirator | 6 | 198 (3.5) | 7.2 (1.2) | 98.6% (0.3%) |

**Table 2:**
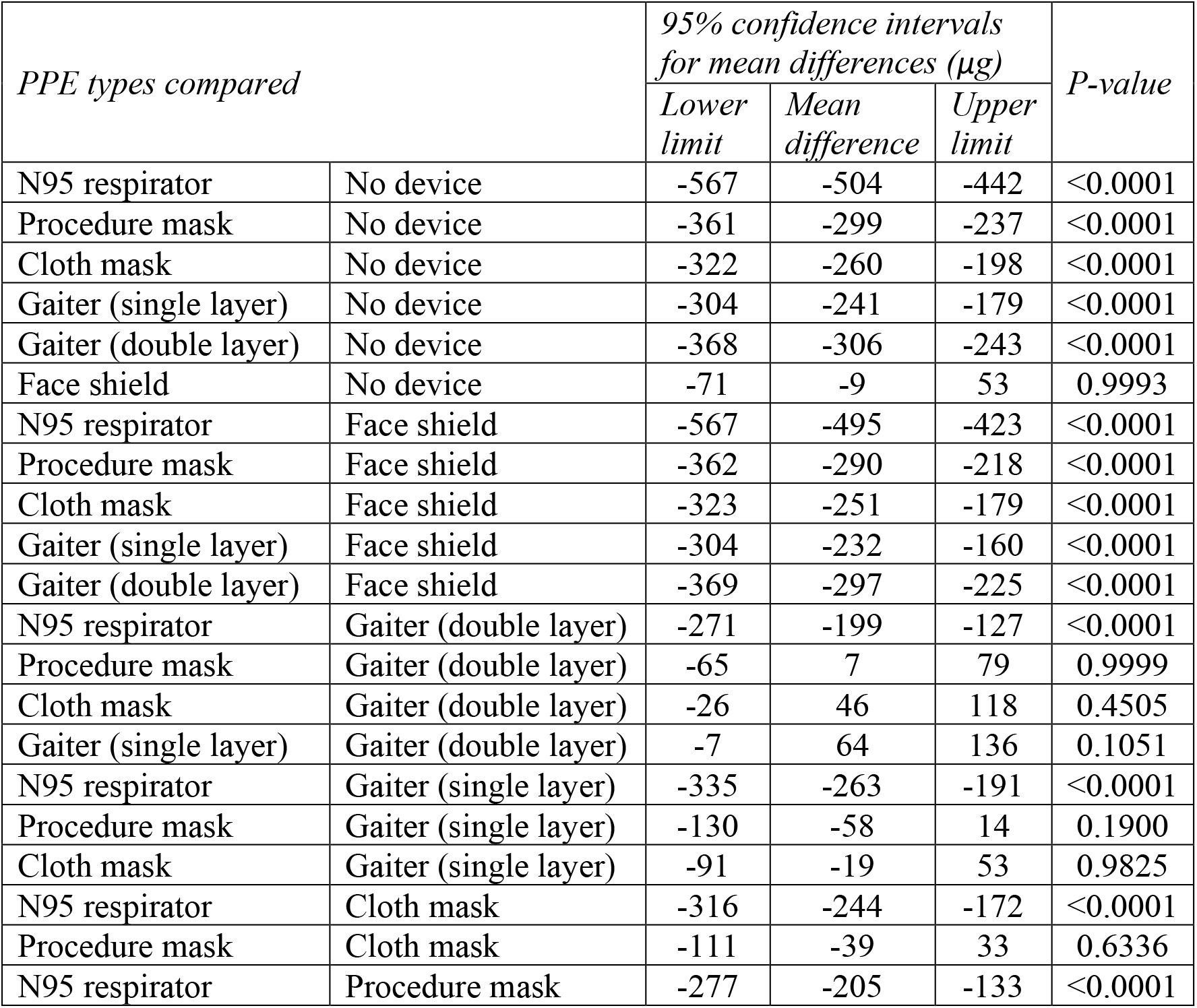
Comparison of devices. Comparison of aerosol mass expelled into the collection chamber while wearing face masks, neck gaiters and face shields.

## Discussion

The amount and sizes of aerosol particles containing SARS-CoV-2 that are expelled by people who are infected are not yet known. Two studies of aerosol samples collected in patient rooms found infectious (replication-competent) SARS-CoV-2 in aerosol particles <4 µm in diameter (Santarpia et al. 2020a) and <10 µm in diameter (Lednicky et al. 2020). Other studies have reported SARS-CoV-2 RNA in exhaled breath from infected patients (Ma et al. 2020), aerosol samples from biocontainment and quarantine units housing SARS-CoV-2 infected persons (Santarpia et al. 2020b), and in aerosol samples at multiple locations throughout two hospitals in Wuhan, China during a COVID-19 outbreak (Liu et al. 2020). The presence of small aerosol particles containing infectious SARS-CoV-2 detected in these studies suggests that in addition to large aerosols, these small aerosols might play a role in SARS-CoV-2 transmission (Anderson et al. 2020; Ma et al. 2020; Morawska and Milton 2020).

Airborne particles larger than 100 µm are ballistic; that is, they are affected primarily by gravity and fall quickly to the ground. Respiratory aerosol particles in this size range tend to deposit within a few meters of the source (Prather et al. 2020). As the aerosol particle diameter decreases from 100 µm, a gradual transition occurs where the settling velocity rapidly decreases and the particles remain airborne for longer times. For example, a 100 µm aerosol particle takes 4 seconds to fall 1 meter in still air, while a 10 µm aerosol particle takes 5.4 minutes and a 1 µm aerosol particle takes 8 hours to settle the same distance (Hinds 1999). Air currents such as plumes of warm air rising from the body can lift these particles and extend the time for which they stay in the air. Thus, small aerosol particles can remain airborne for minutes to hours and can accumulate over time in environments with poor ventilation. Small aerosol particles also are easier to inhale and can travel more deeply into the lungs (Vincent 2005).

Source control devices like face coverings and face shields collect respiratory particles larger than 0.3 µm primarily by impaction and interception of the aerosol particles against the fibers or solid surfaces of the device. Small aerosols require much higher air velocities to deposit by impaction than do larger aerosols, and thus are more difficult to block with source control devices (Hinds 1999; Lindsley 2016). Consequently, small aerosols present the most challenging scenario for testing source control devices since devices that block small aerosol particles would be expected to block larger ones as well. Our results show that face masks and neck gaiters can significantly reduce the expulsion of small respiratory aerosol particles during coughing. This suggests that various types of face coverings can make an important contribution to reducing the quantity of aerosol particles containing SARS-CoV-2 released into the environment by people who are infected. N95 respirators, which are worn for personal protection by healthcare workers and others at highest risk of exposure, are also very effective source control devices. In contrast, the face shield blocked very little of the cough aerosol, indicating that face shields are not effective as source control devices for small respiratory aerosols.

The collection efficiencies of all the devices tested increased as the aerosol particle size increased, and this trend would be expected to continue for larger aerosol particles than were tested here. For example, the collection efficiency of the cloth face mask was 28% for the < 0.6 µm particles and increased to 76% for the 4.7 to 7 µm particles. Similarly, the double-layer gaiter blocked 24% of the < 0.6 µm particles and 76% of the 4.7 to 7 µm particles. These results suggest that cloth face coverings would be effective as source control devices against the large respiratory aerosols that are thought to play an important role in SARS-CoV-2 transmission.

Our study has several limitations. We used a single cough volume, air flow profile, and aerosol size distribution for our studies; these parameters can vary greatly from person to person. We examined the performance of these devices during simulated coughing but not breathing or speaking, which have different air flow rates and aerosol size distributions. Some internal losses of the test aerosol particles likely occurred due to settling or impaction on the surfaces of the collection chamber, which may affect the estimates of the collection efficiencies. We only used a single representative example of each type of device. The shape and composition of face coverings vary widely, and this would be expected to affect the performance of individual devices. Some face masks have exhalation valves or vents which could reduce their efficacy as source control devices. The fit of a particular mask to an individual wearer and compliance in wearing the mask correctly (i.e., over the nose and mouth) also are important factors in how well the mask performs as a source control device. The face shield that we tested has a widely used design, but alternative designs are being marketed that provide greater facial coverage and, in some cases, include fabric skirts between the shield and the face. These alternative face shield designs might perform better as source control devices.

Previous studies have shown that face shields provide eye and facial protection to the wearer from droplets and splashes (Lindsley et al. 2014; Roberge 2016). When a face shield is worn in addition to a face mask, the face shield can also help reduce surface contamination of the mask by large aerosols and reduce the likelihood of hand contamination when the mask is removed or inadvertently touched (Lindsley et al. 2014). Our previous study showed that face shields provide some benefits as personal protective equipment when face masks cannot be worn (Lindsley et al. 2014), but as with all personal protection and source control devices, their limitations must be respected. Our results suggest that face masks and neck gaiters are more effective than face shields as source control devices to reduce the expulsion of respiratory aerosols into the environment as a public health measure to reduce the community transmission of SARS-CoV-2.

## Data Availability

Experimental data is available upon request.

## Acknowledgments

We would like to thank NIOSH machinist Bryan Williamson for manufacturing the parts for the cough simulator. We also would like to thank the NIOSH Morgantown maintenance, security, warehouse and housekeeping departments for their assistance and dedication during the ongoing pandemic. The findings and conclusions in this report are those of the authors and do not necessarily represent the official position of the National Institute for Occupational Safety and Health (NIOSH), US Centers for Disease Control and Prevention (CDC). Mention of any company or product does not constitute endorsement by NIOSH, CDC. This research was funded by the CDC. NIOSH is a part of the CDC.

