## Supplemental materials for "Efficacy of face masks, neck gaiters and face shields for reducing the expulsion of simulated cough-generated aerosols"

### Supplemental online material

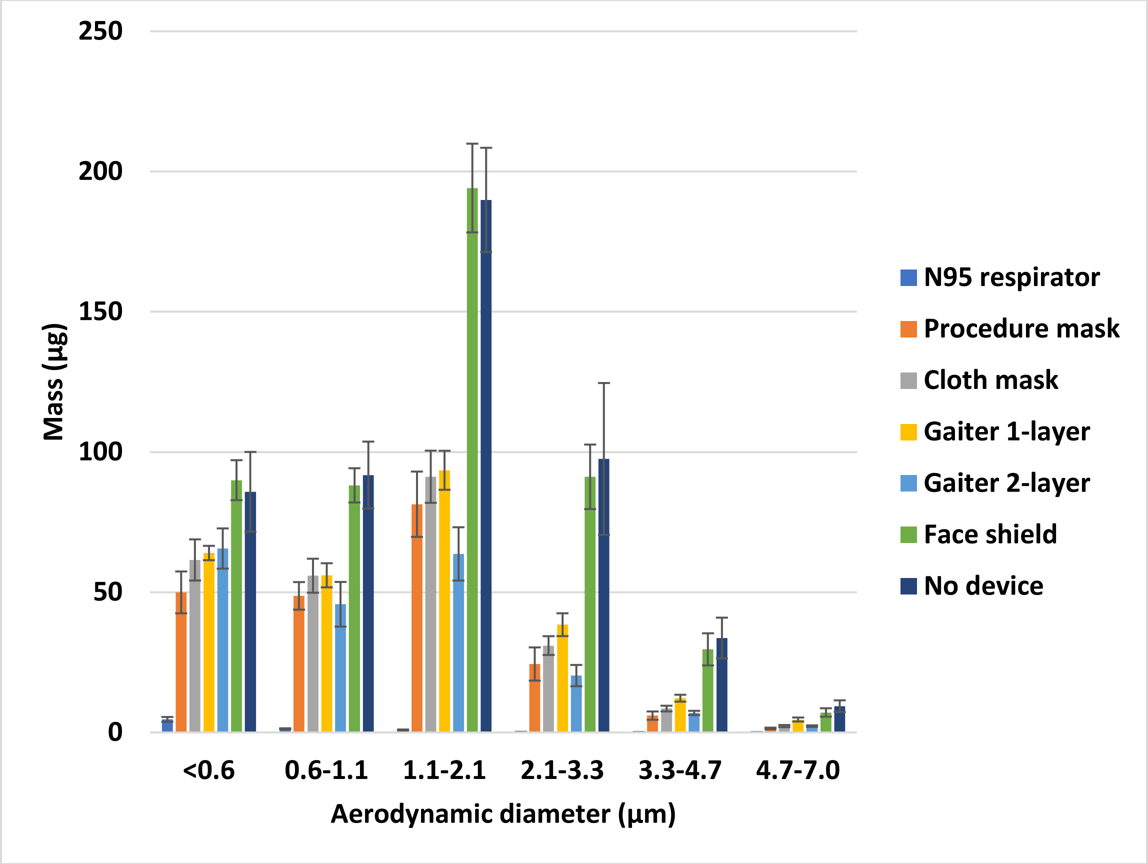


**Figure S1: Color version of Figure 2.** Mass of aerosol collected in each size fraction. The graph shows the amount of simulated respiratory aerosol that was collected from the collection chamber in each aerosol particle size fraction after a single simulated cough. The bars show the mean and standard deviation.

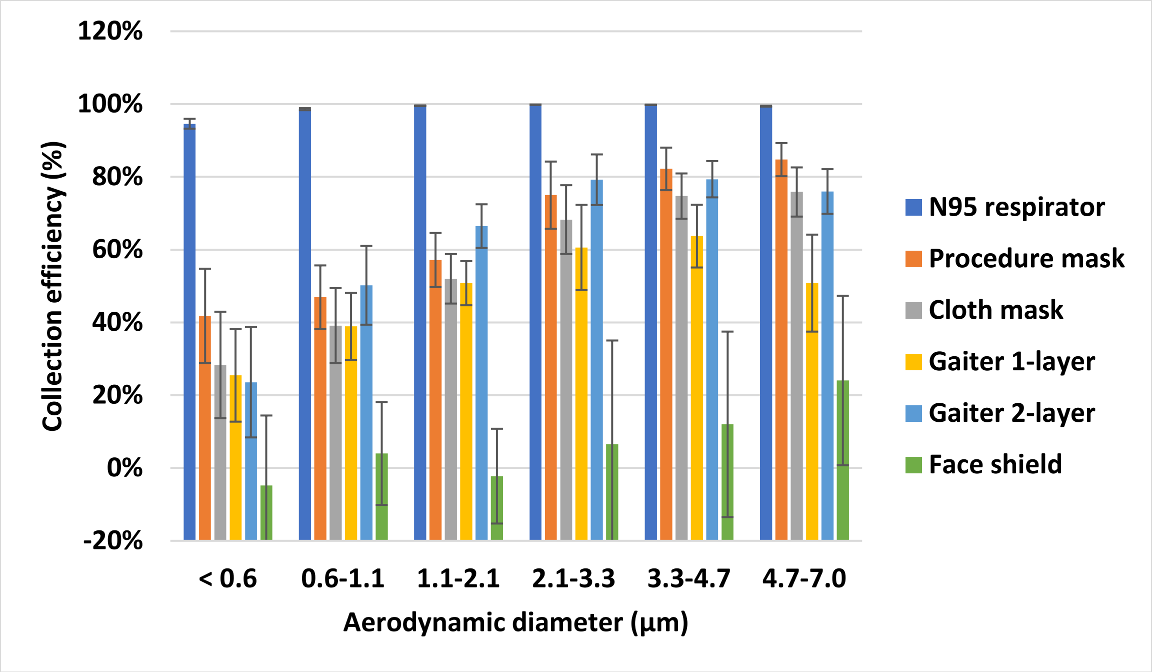


**Figure S2: Color version of Figure 3.** Collection efficiency of face masks, neck gaiter and face shield. The collection efficiency is the percentage of aerosol particles that were blocked by the face mask, neck gaiter or face shield compared with experiments without a device. The plot shows the means and standard deviations of the collection efficiency in each size fraction.

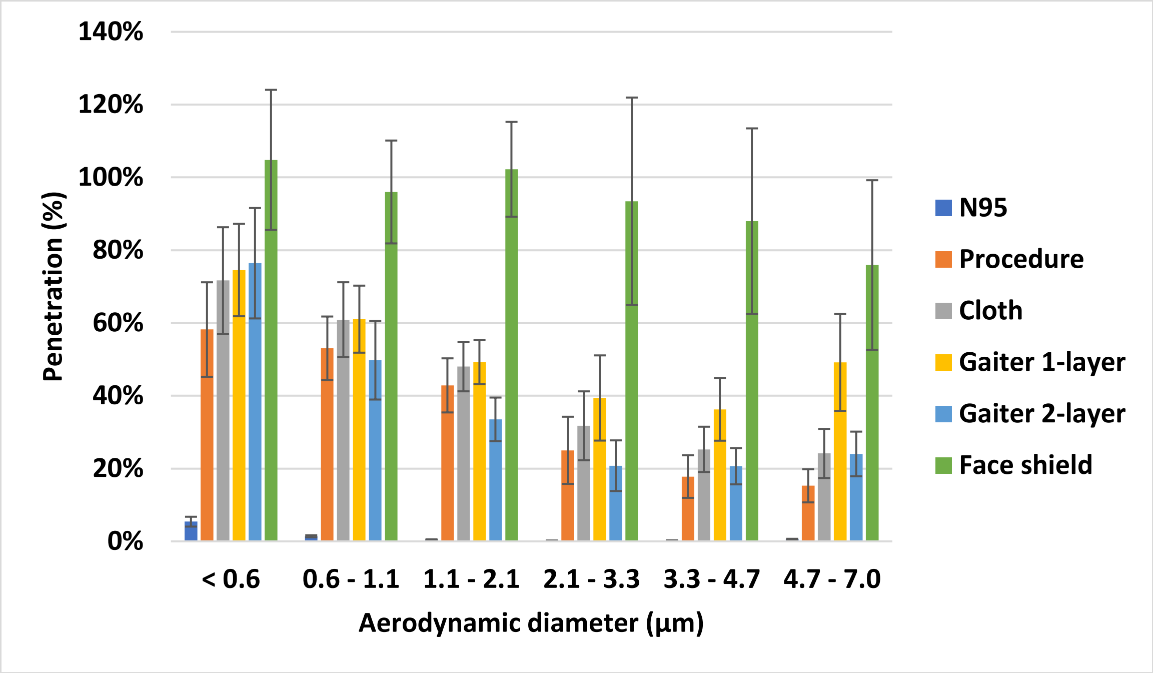


**Figure S3: Particle penetration for different types of face coverings.** This plot shows the same data as in Figure 3, but here it is presented as the percentage of particles that penetrate beyond the face coverings and are collected by the Andersen impactor. The plot shows the means and standard deviations in each size fraction.

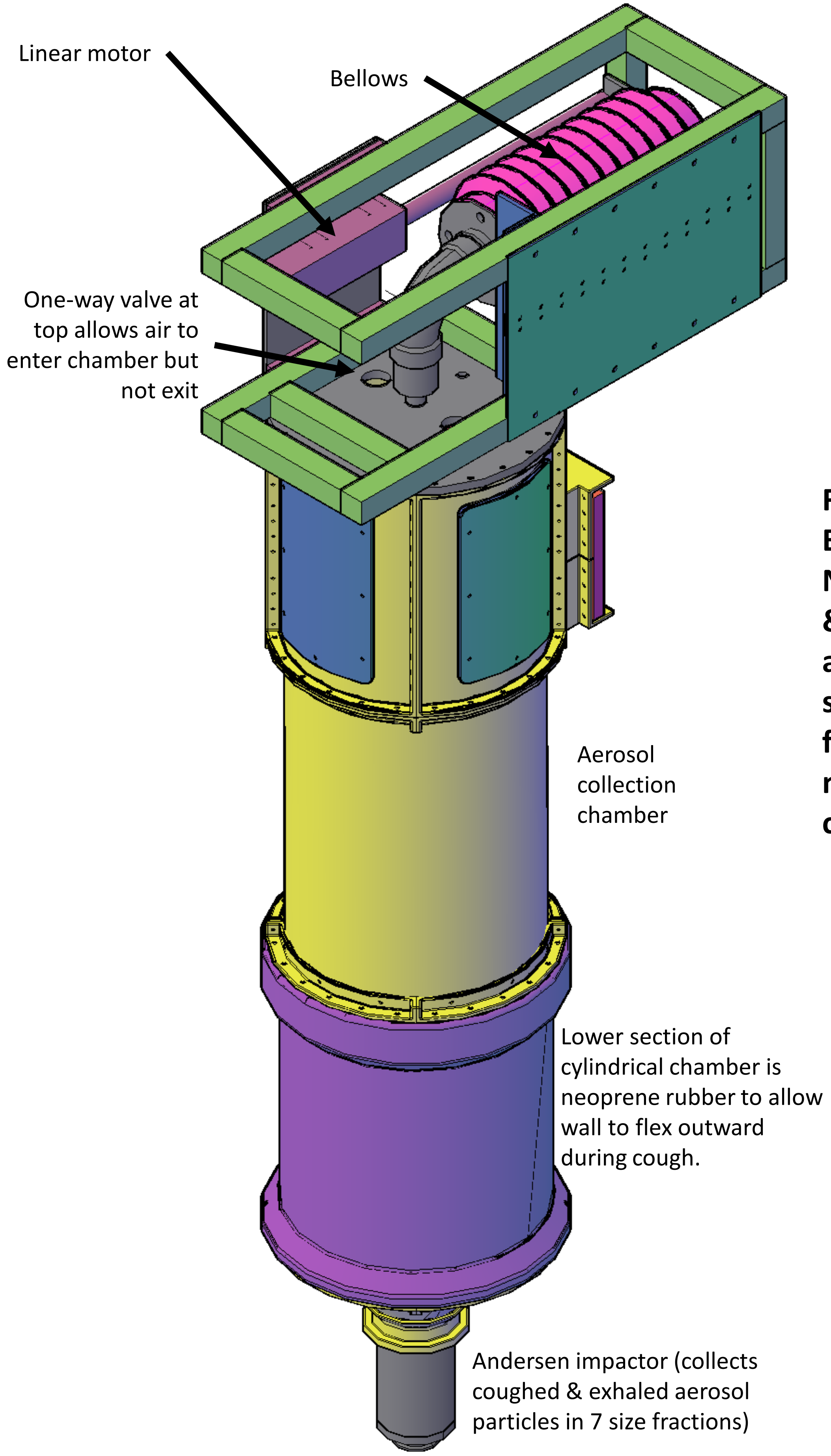


**Figure S4: External view of NIOSH coughing & breathing aerosol simulator for test of masks as source control devices**

**Figure S5: Section view of NIOSH coughing & breathing aerosol simulator for test of masks as source control devices**

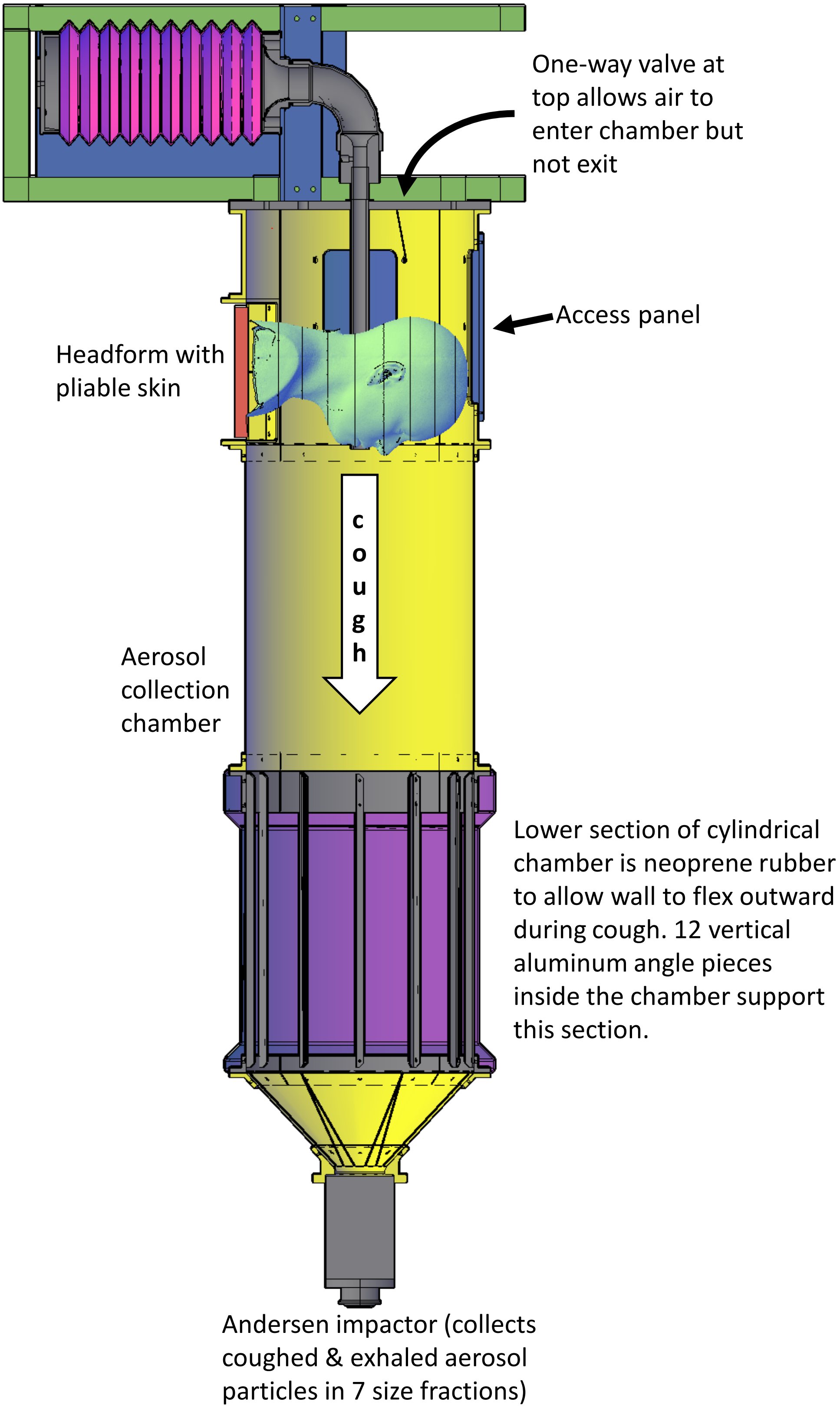

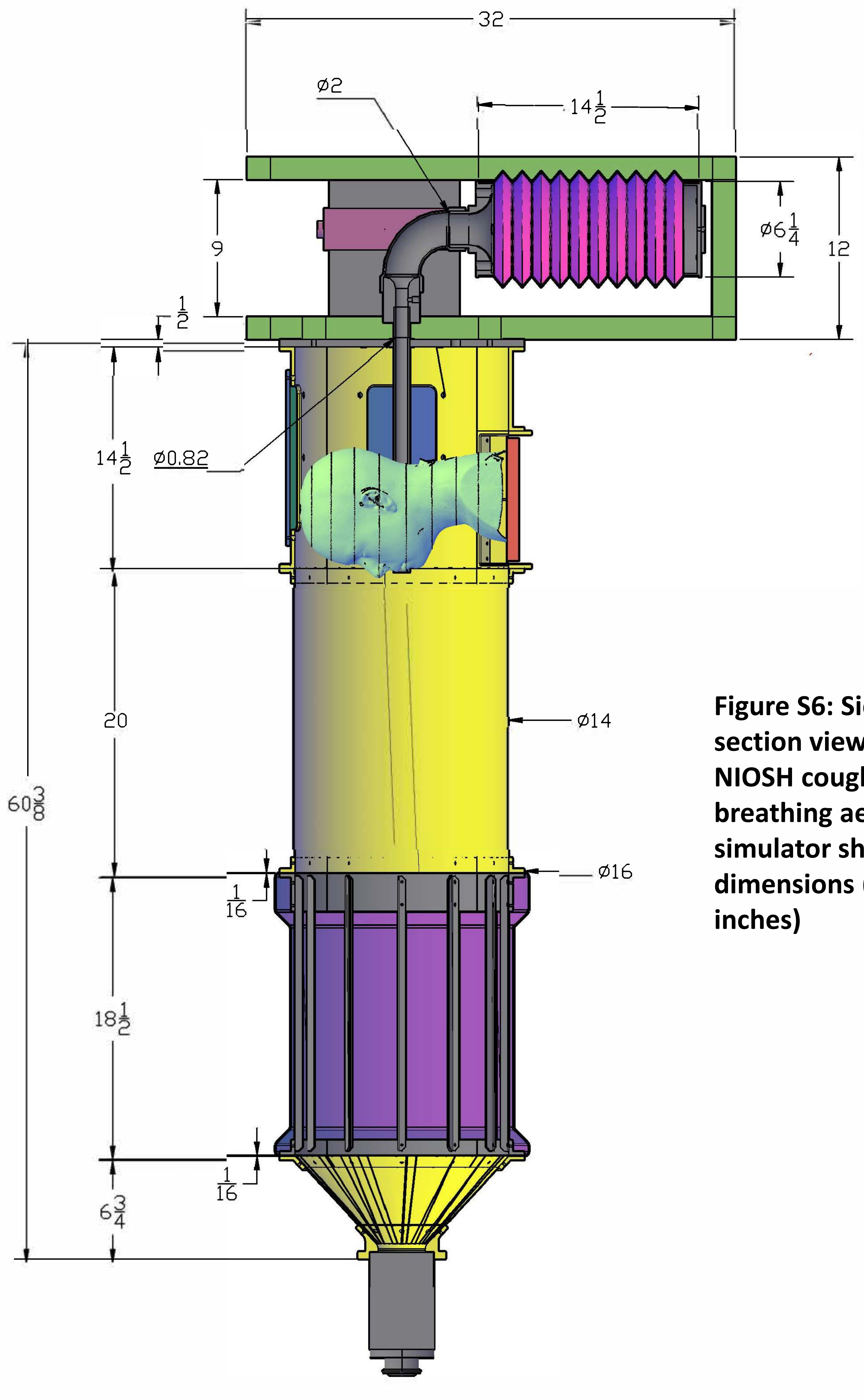


**Figure S6: Side section view of NIOSH coughing & breathing aerosol simulator showing dimensions (in inches)**

**Figure S7: Top section view**

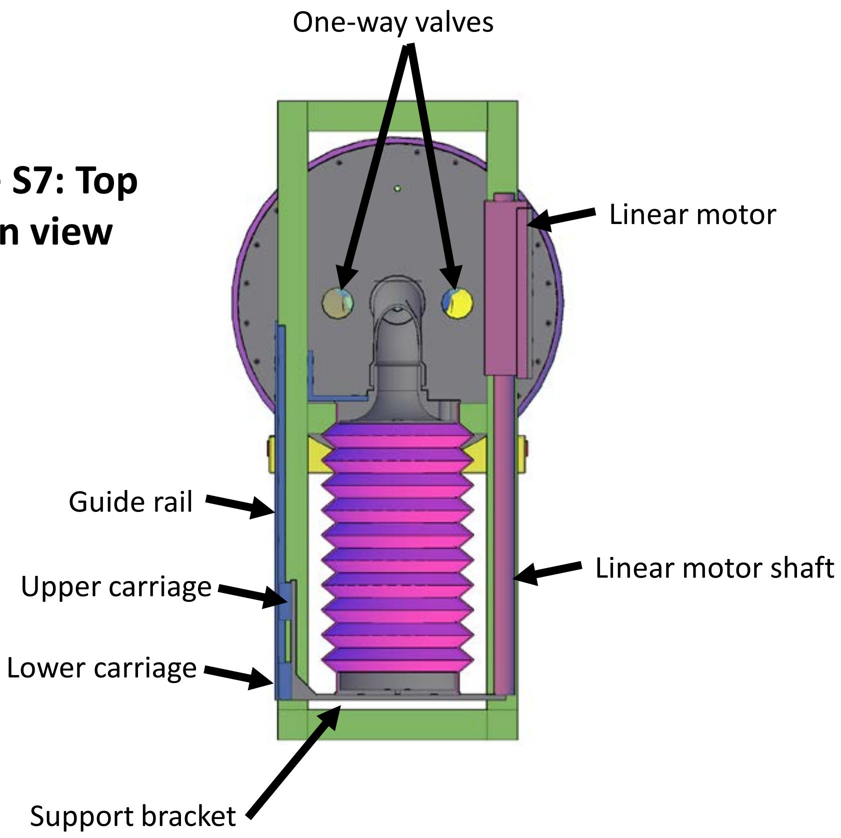


**Figure S8: Bottom view with collection chamber removed**

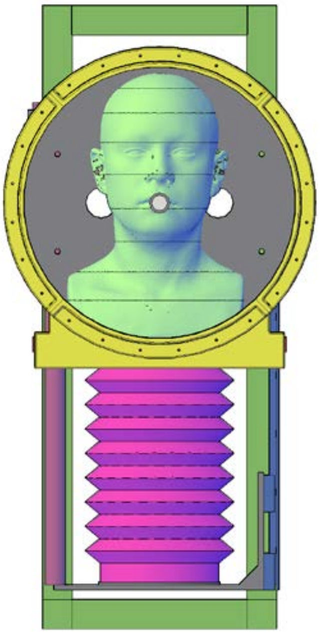

**Figure S9A-F: Pliable skin head form with face coverings**

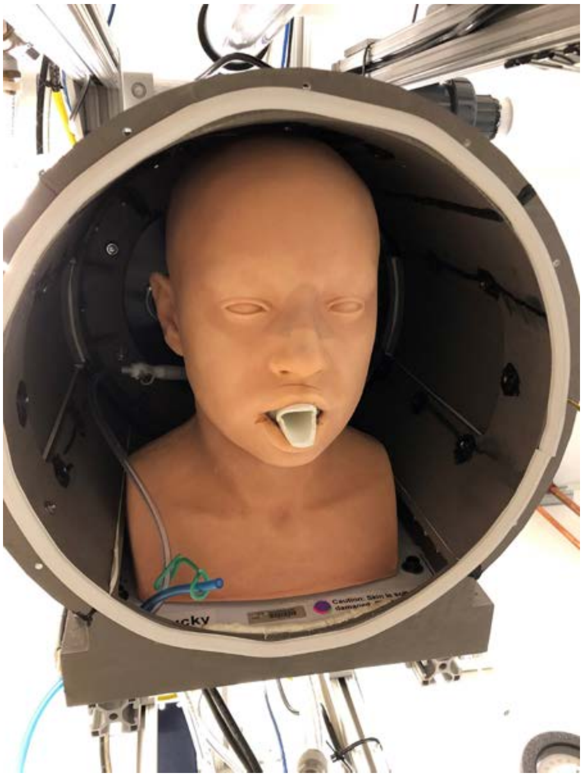

**Pliable skin head form**

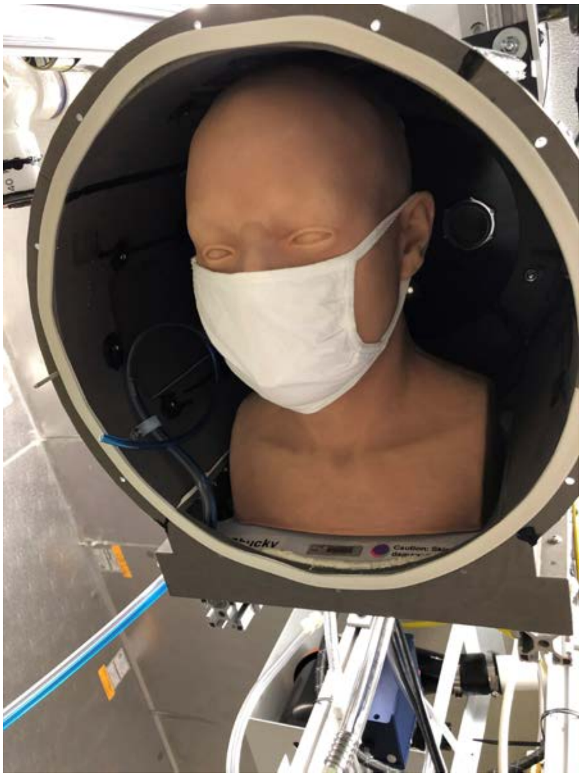

**Hanes cloth mask on head form**

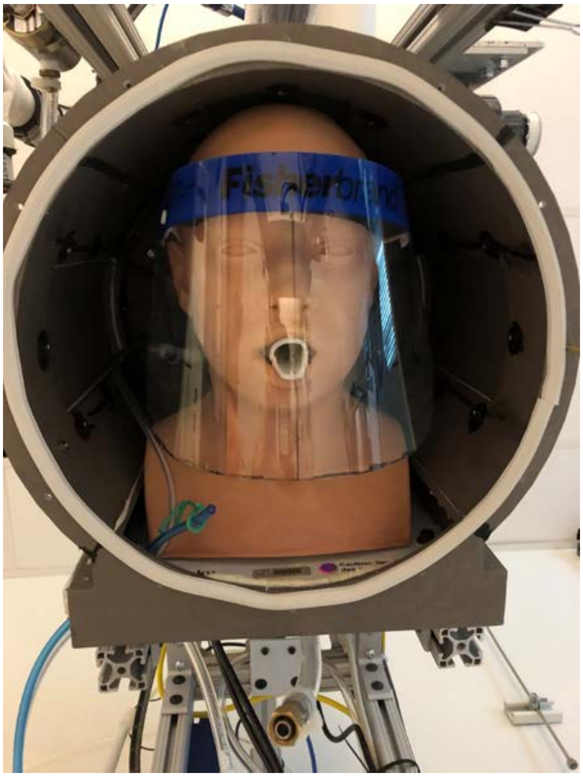

**Fisher face shield on head form**

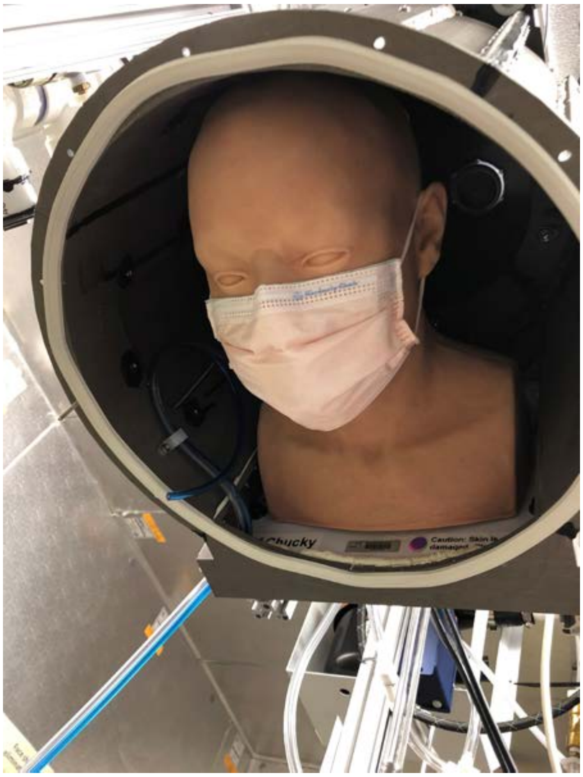

**Kimberly-Clark procedure mask on head form**

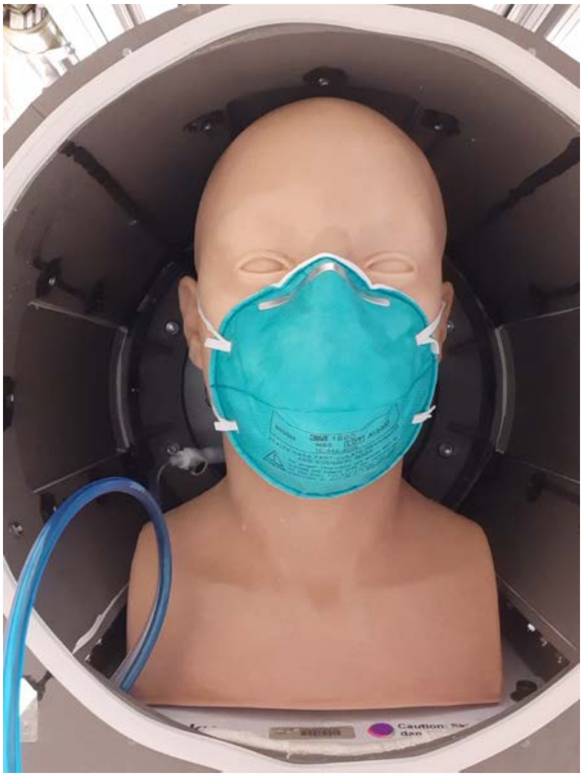

**3M 1860 N95 respirator on head form**

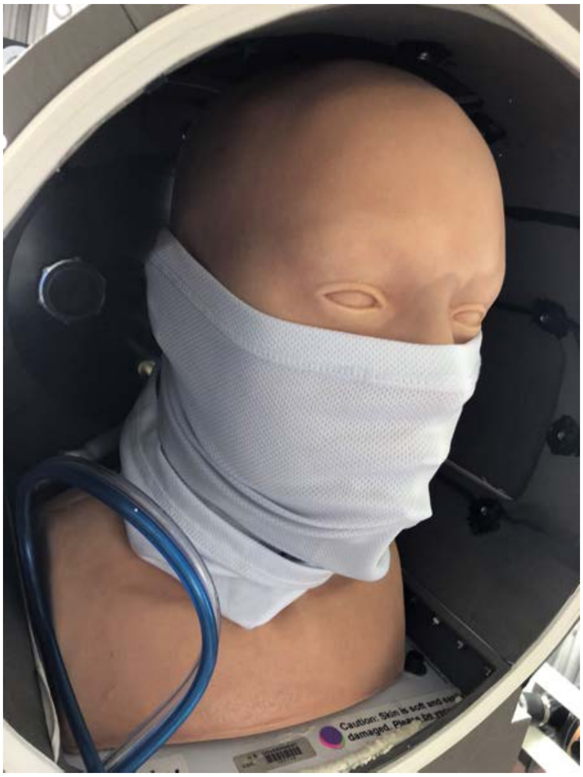

**FKG gaiter on head form**

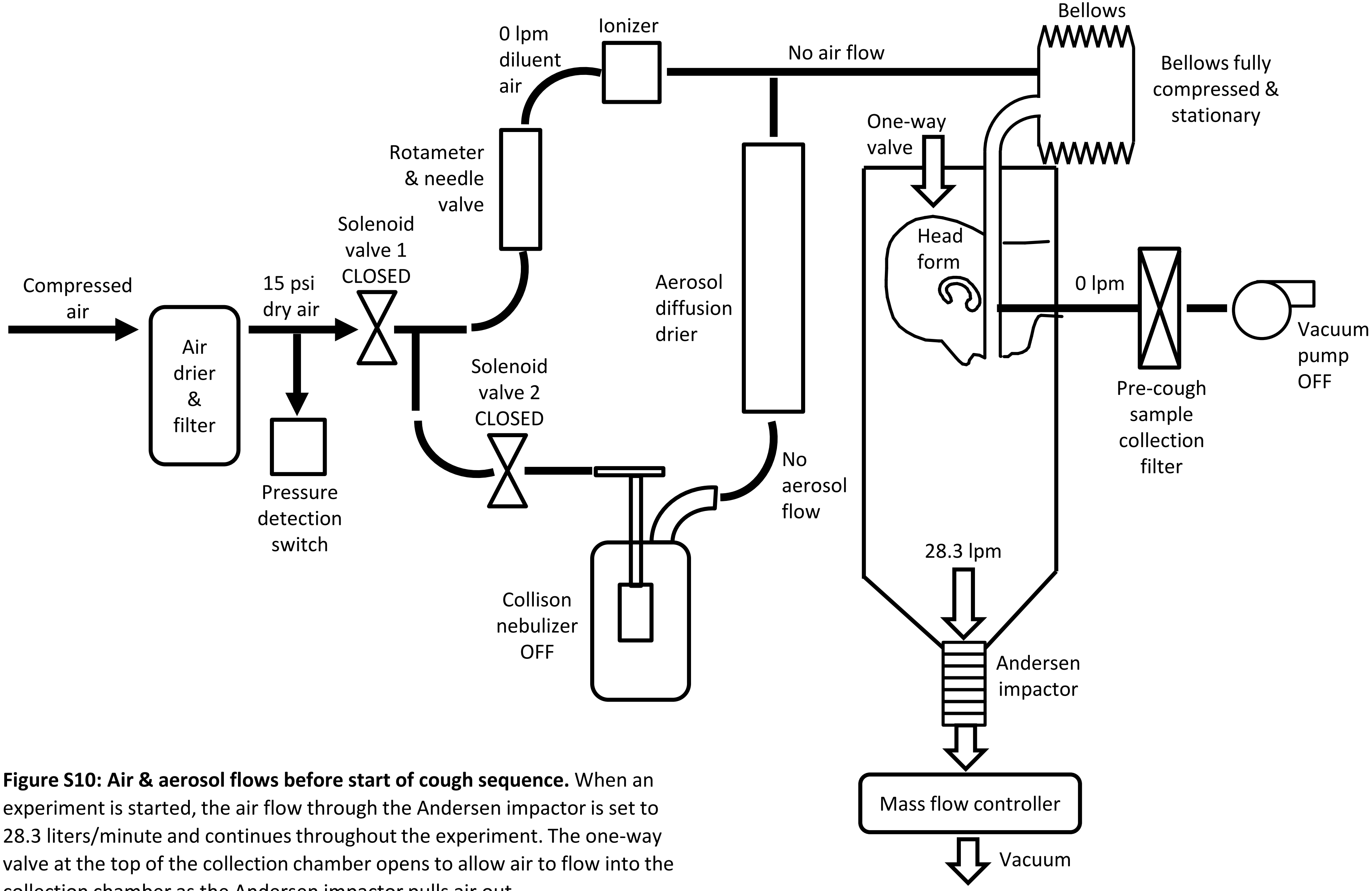


**Figure S10: Air & aerosol flows before start of cough sequence.** When an experiment is started, the air flow through the Andersen impactor is set to 28.3 liters/minute and continues throughout the experiment. The one-way valve at the top of the collection chamber opens to allow air to flow into the collection chamber as the Andersen impactor pulls air out.

The bellows is stationary. Solenoid valves 1 and 2 are closed, so the aerosol generator is not generating an aerosol and air is not flowing into the bellows. Air also is not being pulled through the pre-cough sample collection filter.

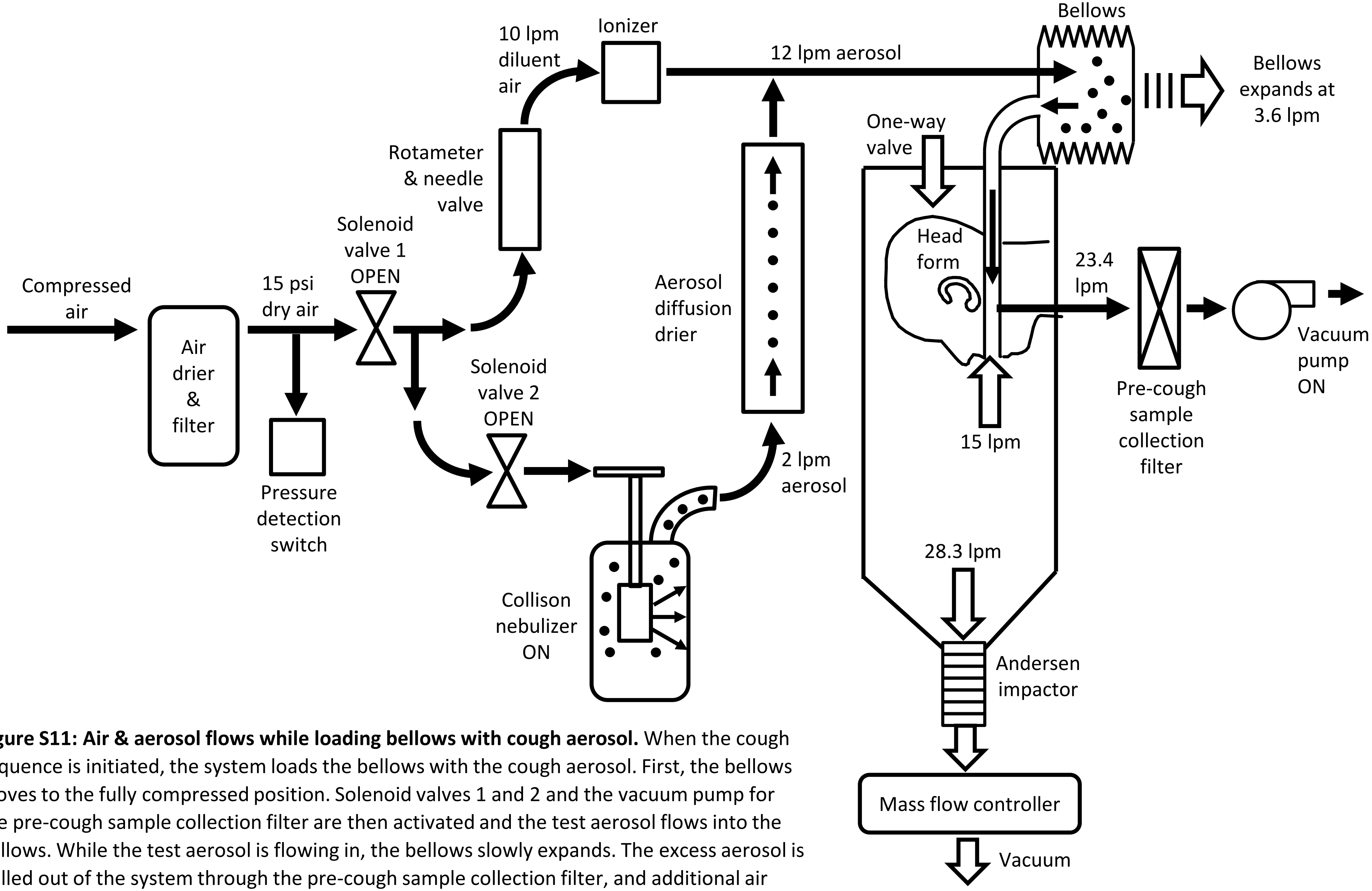


**Figure S11: Air & aerosol flows while loading bellows with cough aerosol.** When the cough sequence is initiated, the system loads the bellows with the cough aerosol. First, the bellows moves to the fully compressed position. Solenoid valves 1 and 2 and the vacuum pump for the pre-cough sample collection filter are then activated and the test aerosol flows into the bellows. While the test aerosol is flowing in, the bellows slowly expands. The excess aerosol is pulled out of the system through the pre-cough sample collection filter, and additional air flows into the mouth from the collection chamber so that aerosol does not flow out of the mouth during the aerosol loading phase. In addition to collecting the excess aerosol, the pre-cough sample collection filter provides a reference sample indicating how much aerosol is in the bellows before coughing relative to other experiments. The Andersen impactor continues to operate during this stage.

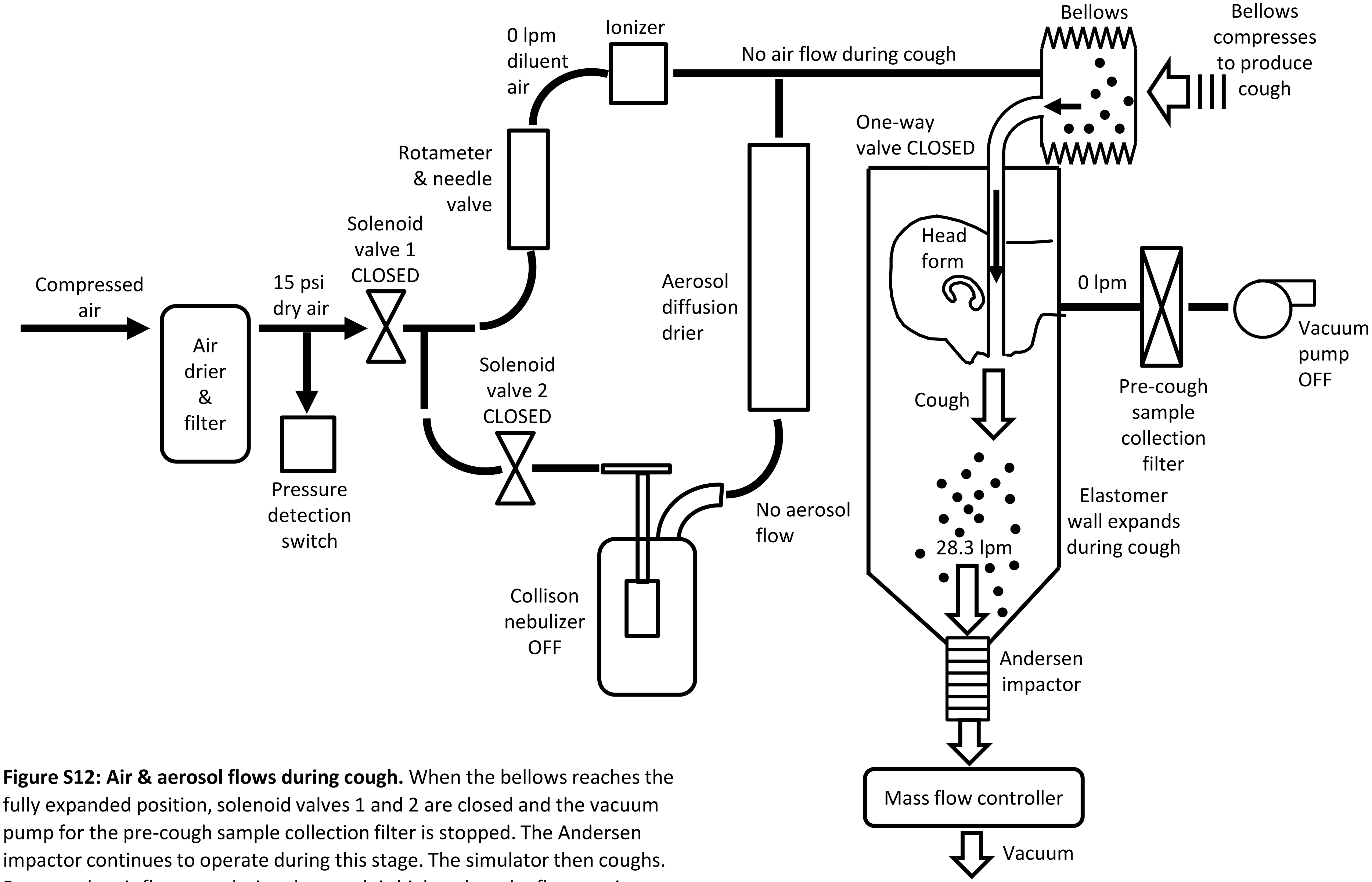


**Figure S12: Air & aerosol flows during cough.** When the bellows reaches the fully expanded position, solenoid valves 1 and 2 are closed and the vacuum pump for the pre-cough sample collection filter is stopped. The Andersen impactor continues to operate during this stage. The simulator then coughs. Because the air flow rate during the cough is higher than the flowrate into the Andersen impactor, the cough causes the one-way valve to close, which prevents the cough aerosol from flowing out of the collection chamber. The elastomer wall in the lower part of the chamber expands to accommodate the volume of the cough.

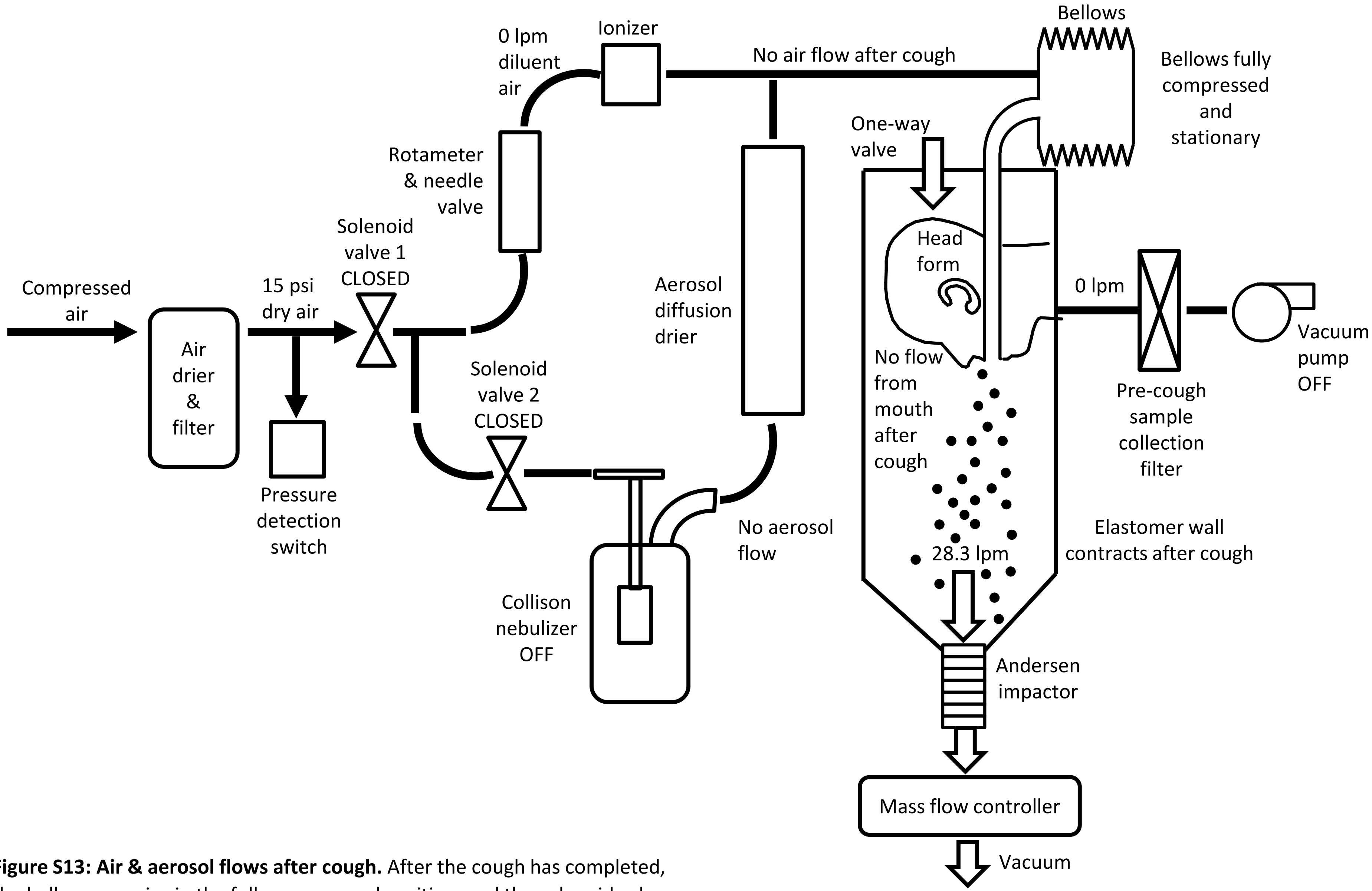


**Figure S13: Air & aerosol flows after cough.** After the cough has completed, the bellows remains in the fully compressed position, and the solenoid valves and the vacuum pump for the pre-cough sample collection filter remain off. The elastomer wall in the lower part of the chamber contracts and the one-way valve at the top of the chamber opens. The Andersen impactor collects the aerosol from the collection chamber for 20 minutes.

**Experimental protocol for test of effectiveness of masks & face shields for source control**

1) If needed, prepare dye stock solution by dissolving 10 g of sodium fluorescein in 80 ml of 0.1 M Tris solution (0.125 g/ml).

2) Add 40 µl of Tween 80 to 40 ml 0.1 M Tris base to get 0.1 M Tris/0.1% Tween 80.

3) Coat impactor plates (one set for each experiment)

    a. Plate coating solution: 5 g Glycerol, 40 ml Ethanol, 1 g Brij 35.

    b. Stir or vortex coating solution before use.

    c. Put six 100 mm plate lids on a level surface.

    d. Deposit 0.5 ml of solution in the center of each plate and allow it to spread by itself—do not tilt the plate. Allowing the solution to spread naturally gives a nice smooth coating, while tilting and rotating gives a blotchy surface.

    e. Allow to dry for at least 30 minutes. Overnight is fine.

4) Check that the silica gel inside the diffusion drier is dry (cobalt blue).

    a. If the silica gel in the diffusion drier has begun to change color to clear, regenerate it or change to fresh batch.

    b. Regenerate silica gel by heating it to 350°F overnight. Keep in mind that the visible silica gel is furthest away from the aerosol and so will be the last to change color.

5) If the diffusion drier is not attached, attach it. Adjust the drier so that it is oriented vertically. Make sure that all Swagelok fittings are tight.

6) Put a 47 mm 5 µm PVC filter in an inline filter holder. Attach the filter holder to the vacuum line (inlet and outlet must be attached to connections). This referred to as the vacuum inline filter.

7) If a mask or face shield are to be tested, place them on the head form.

8) Take a photograph of the PPE on the head form.

9) If testing a face mask, perform a fit test using the PortaCount. See the PortaCount protocol.

    a. **Important:** Make sure that the vacuum inline filter holder is installed and connected to the vacuum pump, or cap the line leading to the inline filter. Otherwise, particles will be drawn into the bellows through the inline filter line and the fit test reading will be too low.

10) Attach the cough aerosol collection chamber to the simulator.

11) Load impactor plates and filter into Andersen impactor.

a. Record the serial number (4 digit) of the Andersen impactor.

b. Place filter holder upright.

c. Place small notched ring in filter holder with notches facing downward (away from support screen).

d. Place the stainless-steel screen in the filter holder with the “inlet” side up (facing the filter).

e. Place a 5 µm PVC filter on top of the screen with collection side up (collection side is up when filter is in package from supplier)

f. Place rubber O-ring on filter holder and press into place

g. Place stage 1 on top of stage 7 to protect O-ring groove, invert, and place on stand.

h. Place filter holder evenly on top of stage 7.

i. Clamp filter holder together

j. Remove stage 1, turn stage 7 right-side up and place in holder

k. Add numbered stages and coated Petri plate lids in order. Be sure to line up the numbers on the sides of the impactor stages.

12) Turn on the power strip for the cough simulator system.

13) Check that mass flow controller has power (display is on).

14) Turn on the Newport photometer. When the display reads “Calibration due”, hit continue.

15) Turn on the Cough simulator control computer.

16) For cough experiments, the vacuum needle valve on the vacuum pump should be completely screwed in. For breathing experiments, the vacuum needle valve should be adjusted to give zero air flow in or out of mouth. See the protocol “Balancing air flow for breathing experiments”.

17) Connect the mass flow control vacuum line to the impactor.

18) Check that vacuum is on (yellow handle & gauge to the right of the Cough simulator computer).

19) Turn on the house compressed air (yellow handle on drier mounted to wall of environmental chamber to the right of the Cough simulator computer). Check that the air pressure is 15 PSI (large gauge on top of grey pressure regulator). You should not need to adjust this.

20) Turn on the TSI flowmeter and connect it and the top of the impactor to the impactor.

21) Use the Cole-Parmer Mass Flow Control program to check the flow rate through the Andersen impactor. Adjust the “desired flowrate” as necessary to get 28.3 liters/min. Note the final setting.

22) Attach the Andersen impactor to the bottom of the cough aerosol collection chamber.

23) On the cough simulator computer, open the program “Bart & Homer check microswitches & pump.vi”.

24) Check that the cough simulator is set to Homer (this is necessary so that the program uses the correct NI-DAQ task ID’s for data I/O, which correspond to the correct NI-DAQ modules).

25) On the Front Panel, start the VI by clicking on the white arrow on the left side of the tool bar.

26) Click on the “Vacuum pump” switch to start the vacuum pump and open the solenoid valve for the compressed air. The “Air pressure” light should come on. If not, stop the vacuum pump, check that the signal cable and all hoses are connected, and try again.

27) Check that the vacuum pump air pressure is 20 PSI (gauge on the vacuum pump). The vacuum needle valve on vacuum pump should be completely screwed in for cough experiments but should be adjusted to give zero air flow in or out of mouth for breathing experiments.

28) Set the diluent air flow (left-hand rotameter) to **10 liters/min for cough experiments or 15 liters/min for breathing experiments.** Flow rate should be read at the bottom of the of rotameter ball.

29) If the Aeroneb nebulizer is being used, check that the flow to the nebulizer is 2 lpm (right-hand rotameter).

30) Click on the “Vacuum pump” switch again to close the valve and press the “stop” button to stop the program. The program will take a few seconds to complete.

31) Before connecting the nebulizer to the diffusion drier:

    a. Add 30 ml of MilliQ H2O to a clean Collison nebulizer.

    b. Connect the Collison nebulizer to a 20 psi compressed air source or to the compressed air source on the cough simulator.

c. Visually check that the nebulizer is producing a strong spray and is not clogged.

d. Empty the nebulizer

32) Attach a clean nebulizer to the diffusion drier and connect the air supply.

33) Add 1 ml of 0.125 g/ml sodium fluorescein stock (1 g/8 ml) to 30 ml of 14% KCl.

34) Add solution to clean 1-jet Collison nebulizer. Check that bottom of nozzle sticks down about 3/8” into fluorescein solution.

35) Turn on the chamber HEPA filter. It can run continuously throughout the experiments.

36) If performing cough tests (as vs. breathing tests):

a. Open the cough machine control program (Cough system controller w mass flow controller.vi).

b. Check that cough volume is set correctly.

c. Check that the cough type is set to Aerosol.

d. Check that bellows type is set to elastomer.

e. Check that the mass flow controller is set to correct value to get 28.3 liters/minute of actual flow (as determined during calibration above).

f. The program will create a log file each time it is run that records when each cough occurs. The file name includes the experiment ID, the date, and the time the file was created.

g. Set the number of coughs to 1.

h. Set time to wait after cough before homing bellows to 20 minutes.

i. If the temperature & humidity sensor is connected, turn on the Collect temperature/humidity data button and set the port to COM10.

j. Check that “Use Andersen Impactor” button is on.

k. Start the cough control program by clicking on the white arrow in the upper left part of the screen.

l. Enter the experiment ID when prompted.

m. Accept the default names for the data files.

n. When the program first starts running, the cough button will be disabled and grayed out until the system checks are completed.

o. When ready to cough, click on the “Cough” button in the cough control program. The computer will take about a minute to go through each cough. A flashing indicator will show that the cough is in process.

p. Check that nebulizer is running correctly.

q. Continue sample collection for at least 20 minutes after the last cough.

r. After collection is complete, stop the cough control program by clicking on “Stop”.

37) If performing breathing tests (as vs. cough tests):

a. Open the breathing control program (Breathing system controller w mass flow controller.vi).

b. Check that the breathing rate is set correctly (usually 15 lpm).

c. Check that “Time to generate aerosol” is set to 60 seconds.

d. Check that “Time to Breathe” is set to 20 minutes.

e. Check that “Collection time after breathing” is set to 5 minutes.

f. Check that the mass flow controller is set to correct value to get 28.3 liters/minute of actual flow (as determined during calibration above).

g. The program will create a log file each time it is run. The file name includes the experiment ID, the date, and the time the file was created.

h. If the temperature & humidity sensor is connected, turn on the Collect temperature/humidity data button and set the port to COM10.

i. Check that “Use Andersen Impactor” button is on.

j. Start the control program by clicking on the white arrow in the upper left part of the screen.

k. Enter the experiment ID when prompted.

l. When the program first starts running, the breathe button will be disabled and grayed out until the system checks are completed.

m. When ready to cough, click on the “Breathe” button in the control program. A flashing indicator will show that breathing is in process.

n. Check that nebulizer is running correctly.

o. Simulator will breathe for the specified time. It will then stop breathing but continue sample collection for the specified time. After collection is complete, the simulator will stop automatically.

38) Remove the nebulizer and place a cap on the diffusion drier.

39) Remove the vacuum inline filter holder and replace it with the vacuum purge adaptor.

40) Remove the Andersen impactor and attach the purge adaptor. Connect the purge adaptor to the mass flow controller.

41) Open and run the purge program.

42) Label six 10 ml glass vials as Plates 1-6.

43) Preparing standards

a. Prepare 1 tube with 8 ml 0.1 M Tris & 8 tubes with 4 ml each. Label these Nebulizer 1-9. When loading the standards onto the plate, use Nebulizer 2-9 for the standards.

b. Take 8 µl aliquot from nebulizer and add to first tube.

c. Take 2 ml from first tube, add to second tube and vortex. Repeat for rest of tubes.

44) Place the filter from Andersen impactor in 150 ml plastic beaker and add 12 ml 0.1 M Tris with 0.1% Tween 80.

45) Place the filter from the vacuum inline filter holder in a 150 ml plastic beaker and add 12 ml 0.1 M Tris with 0.1% Tween 80.

46) Add 3 ml 0.1 M Tris to a vial labeled for vacuum inline filter.

47) Place both beakers on the Belly Dancer and cover. Run at setting 5 for 20 minutes.

48) After 20 minutes, take 1 ml of wash solution from the vacuum inline filter beaker and add to the vial with 3 ml 0.1 M Tris. This 4:1 dilution should be added to the plate for reading the fluorescence.

49) The wash solution from the Andersen impactor filter can be added directly to the plate (no dilution needed).

50) Add 5 ml 0.1 M Tris to each impactor plate. Use cell scraper to stir and scrape surface of plate to recover as much dye as possible. Remove solution & place in labeled vials (Plates 1-6).

51) Add 3 ml 0.1 M Tris to each plate. Use cell scraper to stir and scrape surface of plate to recover as much dye as possible. Remove solution, place in labeled vials (Plates 1-6), and vortex.

52) Place 200 µl of solutions in clear bottom black 96-well plate as shown in plate schematic for fluorescent assay below.

53) Read plate using Spectra Max spectrophotometer

    a. Insert plate into Spectra Max. Do not use the purple adaptor for bottom reads.

    b. Set to read fluorescence endpoint with excitation & emission wavelengths at 485 nm and 525 nm.

    c. Set well pattern to read.

    d. **Read plate from bottom.**

54) Pour excess nebulizer fluid into fluorescein waste bottle. Fill nebulizer reservoir and dump rinse water into waste.

55) To clean the nebulizer:

    a. Thoroughly rinse the nebulizer and glass jar with MilliQ water.

    b. Unscrew the tip of the nebulizer and rinse the inside and outside.

    c. Inspect the O-ring on the nebulizer shaft for tears or cracks.

    d. Screw the tip back on to the nebulizer until it bottoms out.

    e. Fill the nebulizer jar ¾ full with water and screw jar onto nebulizer.

    f. Attach the nebulizer to 20 psi compressed air and run for 10 minutes. While it is running, check for air bubbles at the top of the tip that would indicate that the nebulizer is leaking. If the nebulizer is leaking, mark it and set it aside for repair.

56) Rinse the stages of the Andersen impactor with water. Thoroughly blow water out of stage holes using compressed air.

57) Gently wipe down face of head form with water to remove any fluorescein.
